# pfb_fhir: A utility to extract clinical data systems into a portable format

**DOI:** 10.1101/2023.06.26.23291922

**Authors:** Brian Walsh, Jordan A. Lee, Kyle Ellrott

## Abstract

**Background:** Fast Healthcare Interoperability Resources (FHIR) is a server specification and data model that allows for EHR systems to represent clinical metadata using a consistent API. There is a critical mass of EHR and clinical trial data stored in FHIR based systems. Research analysts can take advantage of existing FHIR tooling for de-identification, pseudonymization, and anonymization. More recently the BiodataCatalyst consortium has proposed the Portable Format for Bioinformatics (PFB) which is a carrier format for describing raw data and the data model in which it is structured, based on an efficient binary format (AVRO). PFB allows an entire cohort of metadata to be loaded into a research data system. Here, we describe an open source utility that will scan FHIR based systems and create PFB based archives.

**Results:** pfb_fhir scans data from FHIR based clinical data systems and converts the data into a self contained PFB file. This utility identifies types, customizations (extensions), and element connections. It then converts all of these components into a graph model compatible for storage in the PFB specification. The structure of the original FHIR system is faithfully reproduced using the PFB schema description system. All records from the system are downloaded, converted and stored as vertices in a graph described by the PFB file. This system has been tested against a number of different FHIR installations, including ones hosted by dbGAP, The Kids First Data Resource and AnVIL.

**Conclusions:** pfb_fhir helps to unlock the potential of EHR and clinical trial data. pfb_fhir allows researchers to easily scan and store FHIR resources and create self contained PFB archives, called FHIR in PFB. These archive files can easily be moved to new data systems, allowing the clinical data to be connected to more complex genomic analysis and data science platforms. The FHIR in PFB archives generated by pfb_fhir have been loaded into data platforms including the Broad’s Terra system, Gen3 based data system, custom graph query engines and Jupyter notebooks. This flexibility will enable genomics investigators to do more integrated genotype to phenotype association analysis using whichever tools suit their line of research.

## Background

Data systems for clinical meta-data are rapidly evolving and must serve multiple communities. On one end of the spectrum are EHR systems that are deployed at enterprise scale in medical institutes and are primarily geared to serving the needs of clinicians. On the other end are systems seen in genomic research projects, such as what are seen in the NIH Cloud Platform Interoperability (NCPI) effort and the Analysis, Visualization, and Informatics Lab-space (AnVIL) project^1^. Research oriented systems are more flexible, allowing for much more exotic data elements to be connected and integrated for analysis. The flexibility comes from researchers needing to define arbitrary tables and columns to house novel data and concepts. These systems are schema agnostic, informaticians can load them with any arbitrary set of metadata.

Fast Healthcare Interoperability Resources (FHIR) is a server specification and data model that allows for EHR systems to represent clinical metadata for data exchange using a consistent API. The original draft of the FHIR specification was published by HL7, a not-for-profit, ANSI-accredited standards developing organization. Released in 2011 the specification has continued to grow as more organizations adopt the standard. The National Institute of Health (NIH) has seen the value of FHIR not only for healthcare records, but also as a formalized schema to enable sharing of clinical and phenotypic data for research. Statements from the Office of Data Science Strategy recognize that FHIR “can accelerate the use of clinical data for research” and thus “NIH encourages funded researchers to explore the use of the FHIR standard to capture and integrate patient-and population-level data from clinical information systems for research purposes and to use it as common structure for sharing research data”

FHIR schemas are defined in an *ImplementationGuide*. ImplementationGuides are a collection of FHIR structures that define how a given community will use FHIR. The ImplementationGuide can tighten or relax the specification and can also create, extend, and remove resources or vocabularies. Several use-cases leverage the ImplementationGuide: web site documentation, FHIR server validation, and the server’s Capability Statement may identify one or more implementation guides that the server supports. As a useful convention, example data and custom terminologies are bundled into ImplementationGuides. ImplementationGuide authors may publish to a registry, for example https://registry.fhir.org/ has 182 registered.

In order to manage the communication of the diverse selection of data systems, the Portable Format for Bioinformatics (PFB) was proposed as a common carrier format. PFB is an Avro-based file that bundles schema, data, ontologies/controlled vocabularies, and pointers to data files in a single, serializable format. It is a meta-format that contains a record of the data’s schema in the header of the document, and then a complete archive of all records. Because a PFB file carries both the data and its schema, the archive file is self-contained, representing a complete snapshot of a data system, and can represent another number of data models. PFB is seen as a way to communicate between different types of data systems. PFB files can be easily moved between systems and has the flexibility for different data models, being neutral, unopinionated in terms of specifying a schema.

The PFB format was designed specifically to enable research analysis, and because of its need to support a variety of research topics, it is schema agnostic. There is no core schema, and every group that utilizes PFB is free to define every data element used in the archive. Freedom to define data schemas results in incoherent data and cross study cohorts that are impossible without significant data engineering. Conversely, clinical EHR systems take a more rigid, static approach. FHIR finds a midpoint between these views by providing a core schema and allowing for user defined extensions. At its core FHIR defines resources for ResearchStudy, ResearchSubject and incorporates comprehensive clinical Patient data with specific sets of fields and structured links between them. Researchers can further customize the schema by adding extensions that are named spaced and can be supported by downstream systems without prior configuration.

The pfb_fhir utility provides a consistent set of utilities and rules for converting structured FHIR records, and the various extensions that may be present in an ImplementationGuide into PFB files. Once these clinically derived results are structured into PFB files, that can be loaded into research analysis platforms, such as the Broad’s Terra system, BioData Catalyst, Jupyter notebooks or the Gen3 platform^2^.

This effort encourages adoption of PFB as a frictionless mechanism for informatics study data. The pfb_fhir utility addresses a key component for enabling the clinical to research data pipeline. It enables data engineers to leverage FHIR’s association with EHR systems, de-identification services, and terminology services while connecting them to complex data science platforms that allow for integrated analysis.

## Implementation

Our utility pfb_fhir processes raw data extracted from a FHIR based EHR endpoint and converts it into PFB. As seen in Figure 1, this tool traverses the FHIR data model, including custom extensions. The resulting data is easily imported into systems that support PFB (such as Terra, Gen3, and Jupyter) and maintains fidelity to the underlying FHIR schema.

**Figure 1:**
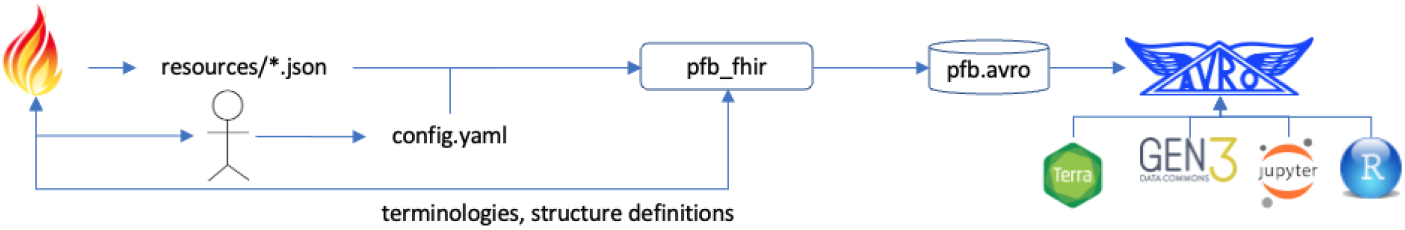
FHIR to PFB data flow. The flow commences post query from a FHIR service, the user provides a config.yaml file to run the pfb_fhir tool which traverses the FHIR data model, including custom extensions ultimately producing a PFB file that can be imported into a downstream system.

While the running of the pfb_fhir utility is primarily automated, there are a number of decision points that need to be configured to tailor data transformation to the user’s needs. The user is expected to describe the intention, the shape, of their extracted FHIR data in a configuration file. FHIR accommodates a multiplicity of optional links between resources. Since the user has queried the data from a FHIR server, the PFB maintains links between nodes. These links are roughly equivalent to FHIR’s _include and _revinclude search parameters that join resources. Thereby, the configuration file describes the actual traversals between nodes (Table 1). Additionally, avro files should be read in dependency order, i.e. a reference to a node must already exist. These answers are captured in a yaml configuration file of resource names in dependency order, each with expected links and an optional structure definition url.

**Table 1:** pfb_fhir Configuration. The user can configure how entity relationships are mapped into the target graph space.

| Entity | Category | Link Name | Link Required | Link Type |
| --- | --- | --- | --- | --- |
| Patient | Administrative | managingOrganization | TRUE | Organization |
|  |  | ⋮ |  |  |
| Condition | Clinical | subject | TRUE | Patient |
|  |  | encounter | FALSE | Encounter |
| Specimen | Biospecimen | subject | TRUE | Patient |

One of the major tasks of the pfb_fhir utility is to examine a FHIR resource’s ImplementationGuide and define conversion steps using the configuration defined by that document. As part of the testing of the pfh_fhir utility, three different ImplementationGuides (Kids First, dbGap and NCPI) have been included in the use case test described in the results section.

During execution the pfb_fhir processes the configuration and dynamically retrieves the FHIR profile and terminologies. The processing pipeline executes a standard set of operations: 1) pfb_fhir reads raw FHIR files that may be formatted as either new line delimited json or FHIR bundles, 2) deserialize into a native python object using python client packages fhir.resources^3^ and smart-on-fhir^4^, 3) map terminologies and extensions, 4) simplify each of the FHIR resources and write FHIR profile attributes to the PFB schema.

From the command line, the utility will simplify, validate and write a PFB file for the 1000 Genomes Dataset, 11 MB in size, consisting of over 22000 FHIR resources in 11 seconds. The graph in the resulting file has 18 different node types with over 19000 edges.

The resulting PFB archive can then be easily imported into the user’s target analysis platform, or accessed directly using software libraries. The pfb_fhir software can easily be installed using the Python Package Index (PyPI). A majority of the program runtime is the FHIR server scraping process, with the actual schema conversion process only taking a few seconds.

### Mapping Challenges

The pfb_fhir utility needs to address several structural differences in order to fully translate FHIR based data into the PFB format. These structural differences have been enumerated in Table 2. The tool accomplishes this in three steps: constrain, simplify and render. Constrain - the user configures what subset of FHIR comprehensive model matches the research domain. Simplify - the tool matches the FHIR nested model to the needs of dataframe like structure of property graphs, indexing and analysis tools. Render - the tool performs a high fidelity serialization of the model and accompanying instance data.

**Table 2:** Mapping issues when translating FHIR to PFB.

| Mapping Issue | User Configurable |
| --- | --- |
| Converting nested documents to structure key-value store | Yes |
| Encoding embedded enumerations and referencing external terminologies | Yes |
| Encoding polymorphic links | Yes |
| Encoding polymorphic values | No |
| Integrating embedded and external schemas | No |

Our destination target, the PFB specification, leverages a property graph style structure where each node is a set of name-value pairs. While nested records are possible within the underlying avro layer, the PFB tooling expects primitive values. PFB defines enumerations in the schema. Edges between PFB nodes are single-typed, with the source-destination node type specified in the schema. Avro defines its own schema structure embedded into each file and presented to avro readers along with the data. This architecture results in compact data representation and supports schema evolution^5^.

This differs from our source, FHIR resources, which are documents with nested resources, extensions and complex data structures that implement a class hierarchy^6^. This document approach is well suited to the domain, as the flexible, semi-structured, and hierarchical nature of documents and document databases allows them to evolve with applications’ needs. As part of the AnVIL project, data from multiple consortiums and over 400 submitters has been harmonized via a high fidelity transformation to the FHIR specification. Additionally, there are a number of FHIR products that can be deployed to start building a compliant data repository^7^. This approach to harmonizing the clinical and research communities is gathering support, for example, the INCLUDE project’s data has been standardized using FHIR for the clinical data and DRS for the genomics data.

Enumerations are defined as *CodeableConcepts*, tied to terminology services, i.e. vocabulary binding [Figure 3]. For example, a practitioner collects a Specimen from a bodySite. FHIR binds the bodySite field to the SNOMED body structure domain^8^. Edges between FHIR resources are multi-typed, the target reference type may be expressed as a list of alternative types, references are expressed in numerous ways (one of *Reference*, canonical or *CodeableReference*). Additionally, FHIR has a somewhat unique mechanism, a polymorphic property, “Choice of Data Types” ^9^, to overload property types. FHIR uses this mechanism to implement a dynamic typing pattern.

One of the most complex steps in the conversion process is how to bridge the gap between FHIR’s document model and PFB’s property graph approach. Specifically, FHIR allows multiple, labeled edges between nodes and nested object structures. PFB is limited to flat key value pairs and a single unlabeled edge from child to parent. The document is simplified into a set of individual graph nodes, with sub objects flattened into the parent, as seen in Figure 2. This step reduces the hierarchy in the resource which can be useful if you want to shape the resource hierarchy into a two dimensional table. Nested documents could be represented in graph space by creating additional node types and relationships. We selected simplification as it reduces complexity in the resulting graph and more naturally fits with the columnar data stores and spreadsheet like interfaces in destination systems.

**Figure 2:**
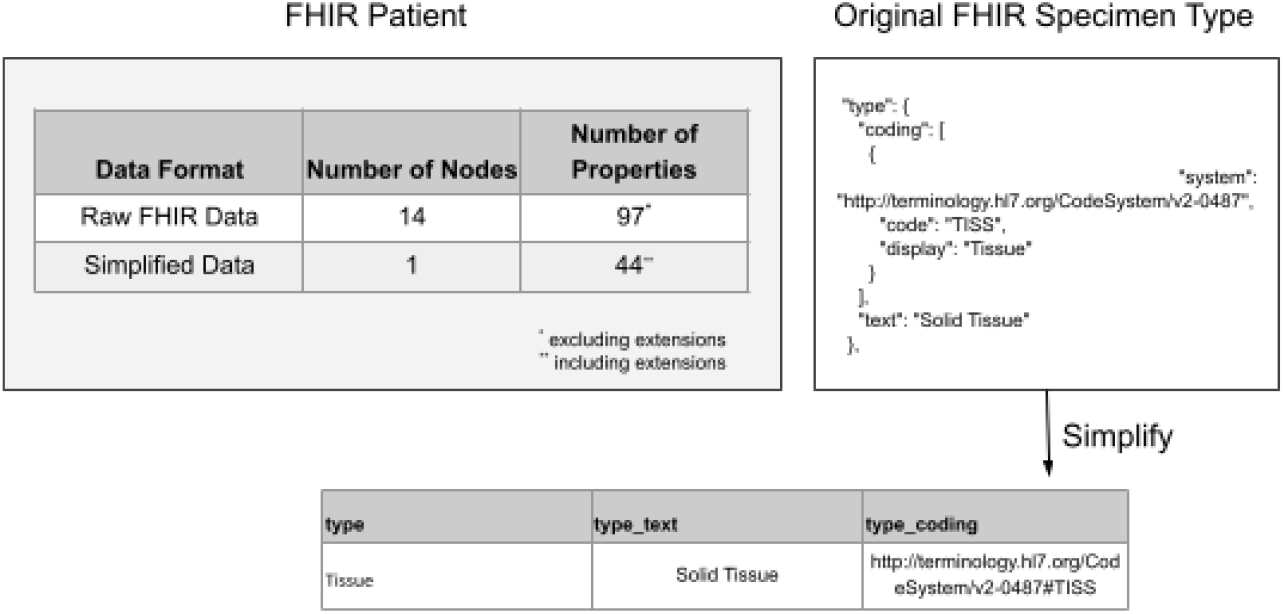
Simplifying Nested Data. FHIR’s verbose description of nested elements can produce output tables that are very cluttered. pfb_fhir has the ability to condense these properties and render uniform datasets that are more analyst friendly.

**Figure 3:**
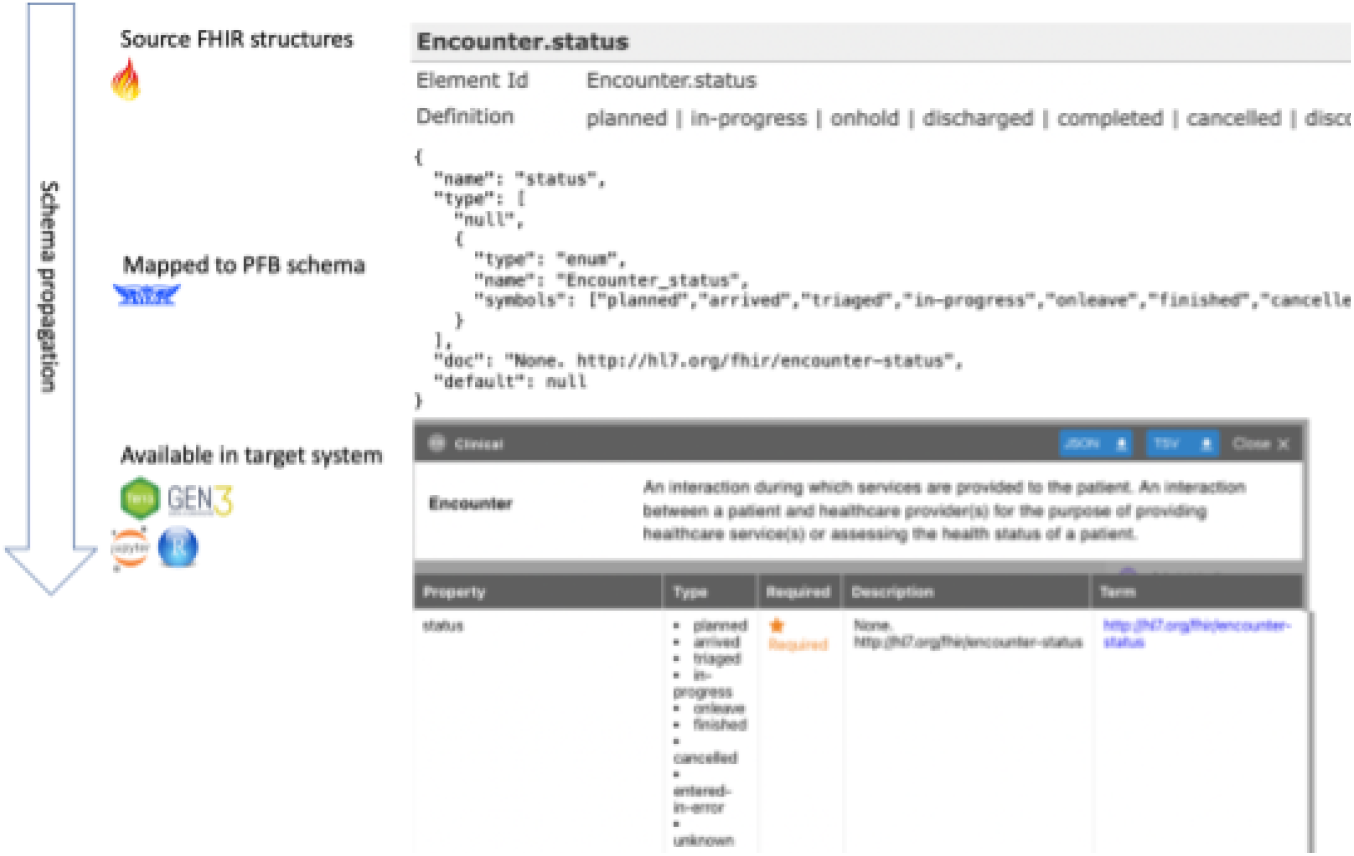
Terminology Linkages. pfb_fhir strives to maintain fidelity to the underlying FHIR CodeableConcept, Coding and Code enumerations. Schema documentation strings are preserved and required bindings are rendered as enumerations.

The design alternative to simply flatten the FHIR resource presented an additional downside challenge, the readability of the resulting data frame in the target tool. For example, a typical Patient resource might have over 70 separate fields in the FHIR resource. Over half of these fields are scaffolding, supporting namespacing, vocabularies and rendering. The result is attribute names with embedded array indices e.g. “identifier_3_type_coding_0_code”. We elected to go with a ‘simplify option’ that will collapse single item arrays, extract coding attributes, collapse extensions and identifiers resulting in a data frame with roughly half the number of attributes. A user can configure the system to extract specific nested properties. The system includes a data utility to simplify FHIR data as well.

As previously mentioned, the schema specifications for each field are transformed into the PFB format. Avro enumerations correspond to FHIR’s *ValueSet*. Specifically, the FHIR binding parameter is used. The binding strength is an indication of the degree of conformance expectations. If the *ValueSet* is bound to a field with strength *required*, or *preferred*, pfb_fhir will generate an enumeration. If the strength is *extensible*, or *example*, a string type will be used. A pointer to the underlying *ValueSet* is published in the schema. A configuration file provided with the software package specifies terminology source files. As fields are discovered, the fhir ValueSet is rendered in the pfb’s *property*.ontology_reference. The user can examine these values by using the pfb utility’s “show metadata” command to render embedded metadata. It is worth noting that in several areas, for example *Condition*.bodySite and *Observation*.code, FHIR suggests values from external ontologies such as SNOWMED and LOINC. These large ontologies with extensible bindings are not rendered as enumerations. The FHIR schema definitions are external to the FHIR data documents. FHIR manages schema changes primarily with extensions and terminology *ValueSets* stored separately. For example, records can have annotated phenotype using the SNOWMED ontology. The structure of SNOWMED, and how the terms are related, is outside the FHIR specification, and annotations stored within FHIR records are simply references to this external standard. Implementers who wish to control these vocabularies may implement additional validation steps.

The PFB defines a link between two nodes with each node defined with a name and id. The name is synonymous with node type. Conversely, a FHIR *Reference* is a field in a resource pointing to another. Depending where it is used in the model, it may be polymorphic, that is it can point to different entity types. Additionally, it may be represented in several ways, as a literal reference, relative reference, or as an internal or absolute url. We use the config file to constrain the destination type.

The PFB (avro), defines a field as a primitive type. While it does support a union type, it does not correspond with FHIR’s “choice[x] type”. FHIR defines these field types as “The value[x] element has an actual name of ‘value’ and then the TitleCased name of one of these defined types, and its contents are those as defined for that type”. For example, the Patient resource structure defines a field ‘deceased[x]’, which indicates if the individual is deceased or not. It’s type is “boolean|dateTime”. In the FHIR json it may be rendered as “deceasedBoolean:” true or “deceasedDateTime”: “2020-07-10 15:00:00.000”. This approach is very similar to object relational mapping’s (ORM) single table inheritance. The utility recognizes these possibilities and correctly renders the actual use in the PFB schema.

One of the core features of Avro is that the schema is embedded in the document. The Avro standard includes support for schema evolution, for example compatibility modes (BACKWARD, FORWARD, FULL,*_TRANSITIVE) and record mutation (field, alias). Similarly, a commonly used set of extensions, us-core is implemented in its own ImplementationGuide^10^. Interoperability is achieved via conformance to the underlying FHIR specification. The pfb_fhir utility captures the schema at the time the data was written, with no attempt made to render differential changes if the schema evolves.

### NIH PFB “light model”

The NIH Systems Interoperation Working Group has defined a “light data model”, a set of fields with commonly accepted meaning [Supplemental Table 1]. We ensure that each entity has a “submitter_id” that defaults to FHIR’s Resource.id or a configurable identifier. This ensures that the original project-specific identifier for this entity is preserved. Additionally, research projects need to connect clinical metadata with large OMICS files such as DIMCOM image archives, and genomic BAM and VCF files. The Global Alliance for Genomics and Health (GA4GH) is a policy group that helps to define technical standards around genomic analysis. The Data Repository Service (DRS) API is a system developed by GA4GH that allows tracking of digital research elements. The DocumentReference.Attachement objects in FHIR can be connected to this API by populating the “gh4gh_drs_uri” field. This allows quick identification of data objects by systems consuming the PFB.

## Results and Discussion

In order to identify utilization of FHIR elements in real datasets, and verify convergence of results, we tested the software on six datasets, and measured the total number of records and edges the results from the conversion. Some concepts, such as Patient and Observations were utilized in every one of the datasets, whereas other concepts, such as Encounter, Immunization, Location and Procedure were only used by one of the datasets. For our analysis, we included datasets from dbGaP, NCPI, AnVIL, Kids First, Genomics Reporting and Synthea. The overview of these data element counts from each of these projects can be seen in Figure 4.

**Figure 4:**
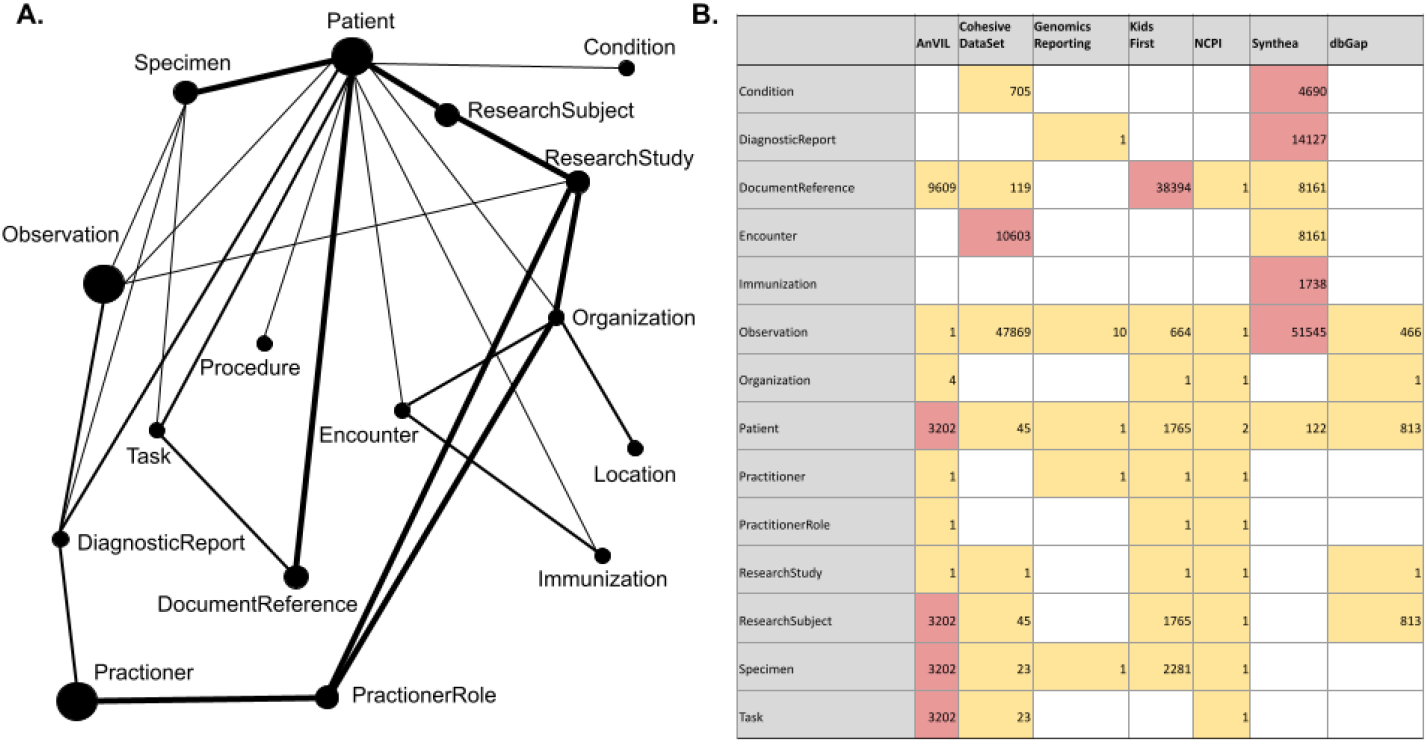
Aggregated demonstration datasets. A) A Graph representation of the unified schema, with vertex sizes representing the relative number of data sets that contain that vertex type and the thickness of the edges representing the number of datasets that implemented that relationship. B) A table of the total vertex type counts across the reported datasets.

The NCBI’s Database of Genotypes and Phenotypes (dbGaP) is one of the data systems currently working to export research phenotypic data using the FHIR specification. The dbGAP data included in the demonstration comes from the synthetic dataset Test Study ALPHA (phs002409). This study was created to simulate an actual patient study for the FHIR API testing project. A single study with 813 patients and 466 observations is included. In this implementation, *ResearchStudy*.extensions defines dbGap variables aggregate study variables such as NumPhenotypeDatasets, NumVariables, and NumSubjects are contained in *ResearchStudy*.extension.

The NIH Cloud Platform Interoperability Effort (NCPI), is a joint effort to connect various NIH data systems, including data from AnVIL, BioData Catalyst, the Cancer Data Research Commons (CRDC), the Kids First Data Resource Center and the National Center for Biotechnology Information (NCBI). By convention, FHIR implementation guides include small example datasets that illustrate extensions and conformance to the specifications defined in the guide. Data for the NCPI is extracted from examples included in the *ImplementationGuide*. A small study with a single Patient is provided. However, all other expected entities are present: *Specimen, Task* and *DocumentReference*. In this implementation study aggregation variables *StorageSize, CohortCount, Participant, SampleCount*, are contained in an *Observation*.*extension*. Additionally, this example illustrates how the utility handles a degree of polymorphism - the *Observation*.focus refers to either a *Patient* or a *ResearchStudy*.

The Kids First Data Resource Center^11^ tracks samples from pediatric research, including childhood cancers and structural birth defects. A single study with 1765 *Patients*, 2281 *Specimens*, 664 *Observations* and 38394 *Document References* are included. The *DocumentReference* url field contains DRS url values. The *Observations* contain *Patient* vital-status values. Although the PFB capabilities weren’t leveraged, the data was used by theNCPI FHIR Code-a-thon to compare tissue-specific gene expression data from open and “registered” access datasets.

Data for the genomic reporting dataset is derived from examples in the *ImplementationGuide*. A small oncology study with a single *Patient* is imported. A complete genomic report including treatment and variants of interest are included in *Observation* and contained by a *DiagnosticReport*. The way this data was serialized as a FHIR Bundle presented some unique challenges. The References between items were coded to refer to the Bundle’s fullURL entries, which in turn are UUIDs separate and apart from the target resource’s id a resource reference can typically be a URI or in this case it can be URI containing a UUID. Additionally, the FHIR specification allows embedding sub-resources known as contained resources^12^ without registering them in the server. These were resolved by pre-processing the data before importing.

We used the Synthea tool^13^, a synthetic patient population simulator to generate data. The configuration was derived from the work done at the GA4GH Computable Cohort Representation Hackathons^14^. The data is a realistic representation of EHR data in a clinical setting, no ResearchStudy exists. Each Patient is connected to a dense set of clinical data: Observation, Conditions, Procedures, etc. Observations also contain questionnaire answers. We also included an enhanced synthea dataset from The “Coherent Data Set”: Combining Patient Data and Imaging in a Comprehensive, Synthetic Health Record^15^.

Data for the 1000 Genomes Dataset^16^ is derived from the work done in the AnVIL project^1^. The data is open access, each patient is connected to a *Specimen* and several *DocumentReference* objects containing urls to cram and bam files. Unlike traditional clinical context, where the data source is derived from a health record system, the 1000 genomes represents anonymized samples that have been represented in the FHIR schema. We additionally tested pfb_fhir’s capabilities for transforming research oriented FHIR data by using it to transform the GTEx dataset^17^ also hosted by the AnVIL project. It is structured similarly to 1000 Genomes dataset, a *ResearchStudy* has many *ResearchSubjects* each with *Specimens* and *Tasks* that contain *Document References* to sequencing files. To validate this dataset and the use of FHIR in research contexts, it was utilized as part of an NCPI FHIR Code-A-Thon^18^, a collaboration with NCPI, Cavatica, and others. To validate the ability to use these datasets in the context of larger integration efforts, the data was harmonized along with data from INCLUDE^19^ and data from Kids First Data Resource Center. Multiple groups have now checked these datasets for Issues around formatting and quality assurance.

### Effectiveness and limitations

Based on the data processed thus far, the utility successfully renders FHIR from a wide variety of use cases. The utility maintains a high degree of fidelity from the source FHIR resources. Given the self contained nature of avro, it should be possible to use the schema as a self contained submission system where the schema checks validity and data integrity. Informaticians could simply write records to the avro file where it would be validated against its embedded schema.

FHIR resources are heavily namespaced and verbose. As such, the resulting data frame is heavily decorated with these urls and enumeration. The simplify option is a first attempt to make the resulting nodes more “data frame friendly”. Additional work will be necessary to reduce FHIR scaffolding. Multiple links can exist between any two FHIR nodes, for example, a FHIR *Task* can have a requestor and an owner that are both *Practitioner* types thereby creating two named edges between the Task and Practitioner nodes. The PFB specification could be improved by adding a label and properties fields to its ‘Link type’ thereby enabling multigraph models.

## Conclusions

We have created a self contained command line utility and API that is able to render a wide variety of FHIR data into the PFB format, making it ready for import and analysis.

We have tested the data against a wide variety of data from different sources, with embedded customizations. The user is required to make a minimal configuration to describe the shape of the incoming data. Run times are reasonable. Future work will decrease the amount of FHIR scaffolding rendered into the PFB nodes, making them increasingly analyst friendly. The roadmap for the system will be driven by collaborator use cases. We will continue to refine and expand the system as we integrate new FHIR instances and PFB consumers. Based on community feedback and use cases, we could add a terminology plug in to provide specific user driven vocabularies such as LOINC and SNOMED, possibly leveraging the Simple Standard for Sharing Ontological Mappings (SSSOM) ^20^ and Google’s Whistle Data Transformation Language^21^.

## Data Availability

All data produced are available online at https://github.com/bmeg/iceberg-schema-tools

## Availability and requirements

Project name: iceberg_tools

Project home page: https://github.com/bmeg/iceberg-schema-tools

Operating system(s): Platform independent

Programming language: Python

Other requirements: None

License: Apache

Any restrictions to use by non-academics: None

### Availability of data and materials

The code and demonstration data sets are available at https://github.com/bmeg/iceberg-schema-tools

### Conflicts of Interest

The authors declare no conflict of interest.

#### Abbreviations

ANSI: American National Standards Institute
API: Application Programming Interface
AVRO: A data serialization framework developed within Apache’s Hadoop project
DRS: GA4GH’s Data Repository Service
EHR: Electronic Health Record
FHIR: Fast Healthcare Interoperability Resources
GA4GH: Global Alliance for Genomic Health
HL7: Health Level Seven
NCPI: NIH Cloud Platform Interoperability
NIH: National Institutes of Health
PFB: Portable Format for Bioinformatics
URI: Uniform Resource Identifier
UUID: Universally Unique Identifier
dbGaP: Database of Genotypes and Phenotypes

### FHIR Entities

Bundle https://build.fhir.org/bundle.html

CodeableConcepts https://build.fhir.org/datatypes.html#CodeableConcept

CodeableReference https://build.fhir.org/references.html#codeablereference

Condition https://build.fhir.org/condition.html

DocumentReference https://build.fhir.org/documentreference.html

ImplementationGuide https://build.fhir.org/implementationguide.html

Observation https://build.fhir.org/observation.html

Patient https://build.fhir.org/patient.html

Reference https://build.fhir.org/references.html#Reference

ResearchStudy https://build.fhir.org/researchstudy.html

Specimen https://build.fhir.org/specimen.html

Task https://build.fhir.org/task.html

ValueSet https://build.fhir.org/valueset.html

## Supplemental Tables

**Supplemental Table 1:**
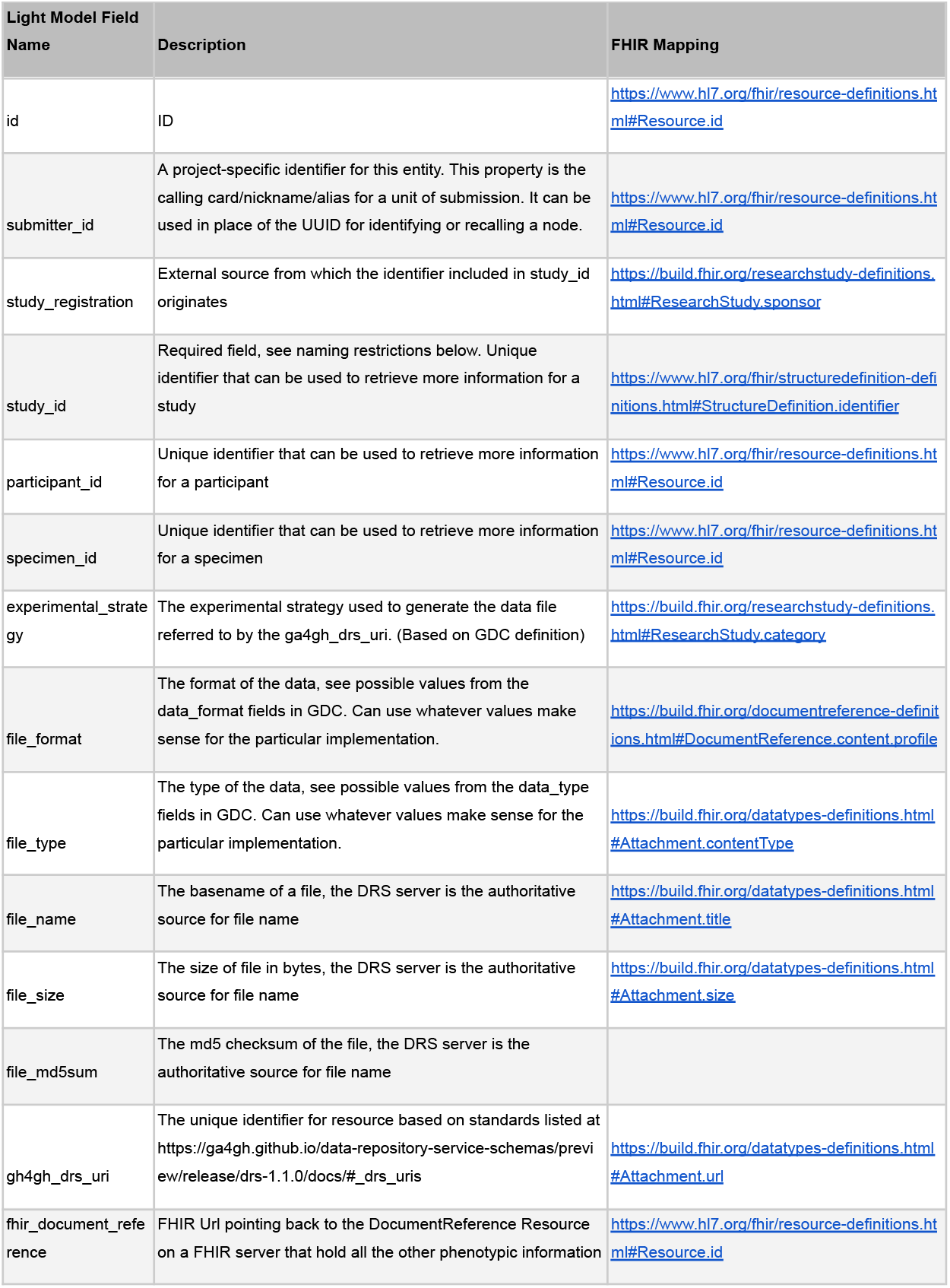
NIH PFB “light model”. A mapping FHIR fields and their mapping to resource types.

